# Chronic Obstructive Pulmonary Disease in adults exposed to fine particles from a coal mine fire

**DOI:** 10.1101/2020.10.14.20213033

**Authors:** Shivonne Prasad, Caroline Gao, Brigitte Borg, Jonathan Broder, David Brown, Jillian Ikin, Annie Makar, Tom McCrabb, Ryan Hoy, Bruce Thompson, Michael J. Abramson

## Abstract

**Introduction:** In 2014 the Hazelwood open cut coal mine burned for six weeks, exposing nearby residents to fine particulate matter (PM_2.5_). The long-term health consequences are being evaluated as part of the Hazelwood Health Study (HHS). These analyses explore the association between PM_2.5_ and chronic obstructive pulmonary disease (COPD).

**Methods:** A sample of 346 exposed, and 173 unexposed, adults participated in the longitudinal Respiratory Stream of the HHS. Participants underwent spirometry and gas transfer measurements and answered validated respiratory questionnaires 3.5-4 years after the fire. Individual level mine fire-related PM_2.5_ exposure was modelled. Multivariate linear regression and logistic models were fitted to analyse associations between mean and peak PM_2.5_ exposure and clinical outcomes, stratified by smoking status.

**Results:** A 10 μg/m^3^ increase in mean PM_2.5_ exposure was associated with a 69% (95%CI: 11% to 158%) increase in odds of spirometry consistent with COPD amongst non-smokers and increased odds of chest tightness (odds ratio; OR 1.30, 95%CI 1.03 to 1.64) and chronic cough (OR 1.24, 95%CI 1.02 to 1.51) in the previous 12 months in all participants. For current smokers, increments in mean PM_2.5_ exposure were associated with higher odds of chronic cough in the preceding 12 months (OR 2.13, 95%CI 1.24 to 3.65).

**Discussion:** Almost four years after a six-week period of coal fire PM_2.5_ exposure, we identified a dose-response association between exposure and COPD in non-smokers. With climate change a likely contributor to increased risk of landscape fires, the findings will inform policy decisions during future sustained smoke events.

**KEY MESSAGES:** *What is the key question?:* Are there long-term impacts of a six-week mine fire event generating PM_2.5_ on COPD and related respiratory symptoms in adults?

*What is the bottom line?:* Almost 4 years after the mine fire, each 10 µg/m^3^ increase in PM_2.5_ exposure was associated with a 69% increase in odds of spirometry consistent with COPD amongst non-smokers, and a 30% increase in odds of chest tightness and 24% increase in odds of chronic cough amongst all participants. Amongst smokers, each 10 µg/m^3^ increase in PM_2.5_ was associated with a 113% increase in odds of chronic cough.

*Why read on?:* With the recent megafires in Australia and the United States exposing communities to smoke for weeks to months, evidence of the long-term health impacts of similar duration PM_2.5_ generating pollution events are needed to inform the public health response.

## INTRODUCTION

The industrial era, largely fuelled by coal, has seen an increase in fires occurring within coal seams and at coal mines in association with human activity and changes in climate patterns.[1] Prominent examples of coal mine fires occurring in close proximity to human settlements have raised concerns about risks to human health around the world.[2,3] Fires at the Jharia Coalfield in India and under the town of Centralia in Pennsylvania, illustrate instances where coal mine fires have led to mass relocation of residents arising from health and environmental effect concerns.[1,4]

The Centralia coal mine fire started in 1962 after local officials set fire to refuse in an abandoned coal pit. The fire then spread along an underground seam to tunnels beneath Centralia.[5] Emission of noxious gases and surface destabilization eventually made the town uninhabitable, with most residents evacuating between 1985 and 1991.[1,5] A health study comparing local residents with those of a nearby town suggested an increase in reported respiratory diseases in men and residents aged 40-79 years relative to their comparators.[6] Two years later a lower rate of diagnosed respiratory disease in relocated residents suggesting some attenuation of effects after exposure to the fire ceased.[7]

Whilst coal mine fires near communities are widely reported, there remains limited peer reviewed literature on human health effects.[1-3] Analogous emissions from wildfires provide some insights into likely effects of exposure to smoke from coal mine fires.[1] Many of these studies assessed exposure to fine particulate matter with an aerodynamic diameter of less than 2.5 µm (PM_2.5_).[8] A review of the health impacts of wildfire smoke in 2016 identified consistent evidence of smoke related respiratory morbidity, particularly asthma and chronic obstructive pulmonary disease (COPD).[8] Wildfire smoke exposure was associated with COPD related physician visits,[9,10] emergency department presentations,[11,12] hospitalisations[13,14] as well as medication dispensing.[9,15] However, almost all of the existing studies used administrative health records to evaluate short term smoke exposure effects. Clinical testing, including lung function measurements, to ascertain effects of bushfires or coal mine fire exposure was lacking.

In 2014, prolonged air pollution was generated from a fire at the Hazelwood open-cut brown coal mine in the Latrobe Valley in south-eastern Australia. Concerns were raised regarding the health of local residents, particularly in the nearby town of Morwell where smoke was visible for 6 weeks. The Hazelwood Health Study (HHS) was established to investigate potential longitudinal health outcomes in people who were exposed to smoke from the Hazelwood mine fire (www.hazelwoodhealthstudy.org.au). The HHS Hazelinks Stream which utilises administrative health datasets, has previously reported more COPD related emergency department presentations,[16] medications dispensed for respiratory condition[17] as well as visits to specialist respiratory services[18] during the mine fire period. The HHS Adult Survey, carried out 2.5 years after the mine fire, also identified higher risks of self-reported respiratory symptoms associated with individual-level mine fire PM_2.5_ exposure.[19] The aim of this further research was to evaluate clinical respiratory outcomes more than 3.5 years after the fire, particularly the risk of COPD and related respiratory symptoms, and their association with individual-level coalmine fire PM_2.5_ exposure.

## Methods

### Study design and setting

This analysis examined cross-sectional data from the Hazelwood Health Study’s Respiratory Stream, a longitudinal follow-up sub-study of the Adult Survey (see the methodology outlined elsewhere).[20] Clinical testing was conducted in Morwell (exposed) between August and December 2017, and in Sale (unexposed) between January and March 2018. Study data were collected and managed using REDCap (Research Electronic Data Capture)[21] electronic data capture tools hosted at Monash University (Victoria, Australia).

### Participants

The Respiratory Stream participants were drawn from the Adult Survey cohort which comprised residents of Morwell or Sale who were aged at least 18 years at the time of the mine fire.[20] Adult Survey cohort members were excluded from eligibility to participate in the Respiratory Stream if they had specified no further contact, had unknown age, sex or were aged over 90 years. Potential participants were also excluded if they had a contraindication to spirometry, such as recent surgery, myocardial infarction, pneumothorax, pulmonary embolus, open pulmonary tuberculosis or known aneurysms.[22] A target sample size of 339 from Morwell and 170 from Sale was derived based on the ability to detect a 5ml/year or greater FEV_1_ decline in exposed compared with non-exposed residents using a two-sample t-test with a two-sided p-value of 0.05 and 80% power. A weighted random sample (to correct for lower response rate in some subgroups of participants, such as young people) of 1,346 Adult Survey cohort members were selected for invitation into the Respiratory Stream, with those who had reported an asthma attack or taking asthma medication oversampled (40%) for further evaluation in an asthmatic sample (see Figure 1). Invitation to participate was by mail, email and/or SMS, and recruitment continued until the target sample size was achieved.

**Figure 1.**
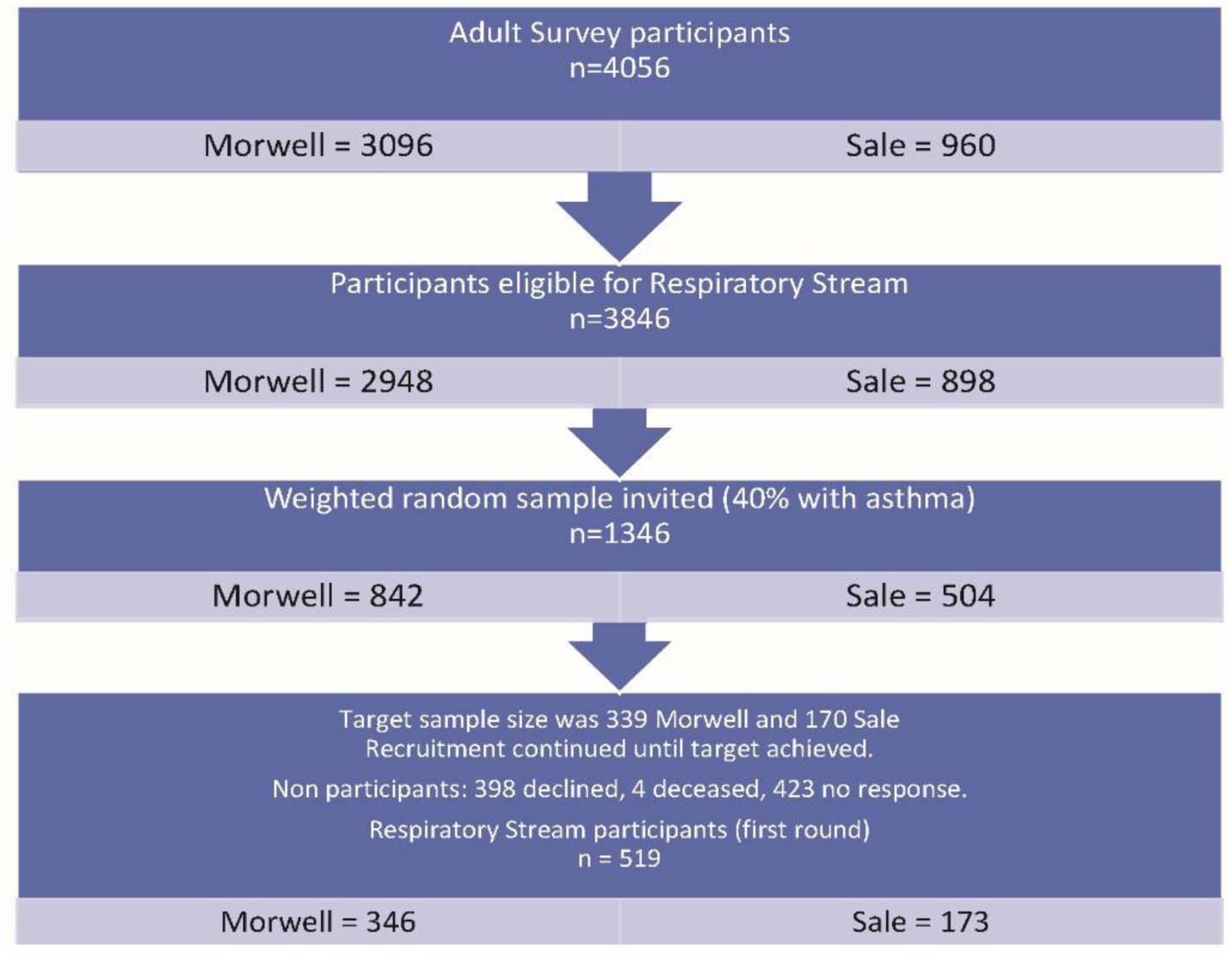
Participant recruitment flow diagram

### Participant characteristics

For each participant, demographic characteristics, such as age, sex, ethnicity, employment status, educational background and occupational exposure (working in dusty or polluted environments, such as coal mines, farms or driving diesel trucks for at least six months) were drawn from the previously completed Adult Survey.[20] Physical characteristics, including height and weight, were measured by trained staff during clinical testing, and body mass index (BMI) was calculated.[23] Smoking history was taken and participants were classified as current smokers, ex-smokers (current non-smokers with >100 cigarettes in their lifetimes) or non-smokers (< 100 cigarettes in their lifetimes).[24]

### Exposure

Due to the lack of ground-level air pollution monitoring from the start of the fire, mine fire-related PM_2.5_ concentrations were retrospectively modelled by the Australian Commonwealth Scientific and Industrial Research Organisation (CSIRO) Oceans & Atmosphere using a chemical transport model that incorporated information on air monitoring, coal combustion and weather conditions.[25] The modelled 12-hourly concentrations were mapped to 12-hourly time-location diaries completed by participants as part of the Adult Survey, to estimate the level of exposure to mine-fire related PM_2.5_ for each individual. Two exposure metrics were considered: mean and peak daily mine fire-related PM_2.5_ exposure. Mean exposure was obtained by averaging the cumulative PM_2.5_ exposure across all the locations that participants visited over the exposure period (9 February - 31 March 2014). Peak exposure was obtained by assigning the highest 12-hourly PM_2.5_ level from all locations, regardless of the time that the participant spent in that location.

### Outcomes

#### Respiratory symptoms and conditions

Self-reported respiratory symptoms and conditions were assessed with standardised questionnaires, adapted from the European Community Respiratory Health Survey II (ECRHSII)[26] and administered by trained interviewers.

#### Respiratory function tests

Trained investigators conducted objective measurements of respiratory function. Standardisation was achieved through the use of Standard Operating Procedures (SOPs), validated equipment, temperature-controlled rooms and utilisation of the same staff and equipment at both clinic sites. Preparation instructions were provided to participants at the time of booking the clinic visit.

Respiratory function was measured using the EasyOne Pro^™^ LAB Respiratory Analysis System (nnd Medical Technologies AG, Zürich, Switzerland). Spirometry was performed according to American Thoracic Society/European Respiratory Society (ATS/ERS) guidelines.[27] Parameters measured included FEV_1_, FVC and FEV_1_/FVC. Z-scores for FEV_1_, FVC and FEV_1_/FVC were calculated using the Global Lung Initiative (GLI) 2012 spirometry reference equations.[28] This analysis focussed on spirometry measurements performed ten minutes after administration of 300 µg Salbutamol via pressurised Metered Dose Inhaler and spacer (pMDI).[29]

Carbon monoxide transfer factor (T_L_co) was measured using the single-breath testing technique according to ATS/ERS guidelines.[30] T_L_co values were corrected for haemoglobin (Hb) using standard equations from ATS/ERS guidelines.[30] Z-scores for T_L_co were calculated using the GLI 2017 reference equations.[31]

### Definition of COPD and abnormal T_L_co

COPD was defined by post bronchodilator FEV_1_/FVC <5th percentile of predicted (lower limit of normal with FEV_1_/FVC z-scores<-1.645) and abnormal T_L_co was defined as haemoglobin corrected T_L_co <5th percentile of predicted (T_L_co z-scores<-1.645). A similar definition for COPD was used in a recent population-based study on air pollution, lung function and COPD.[32]

### Statistical methods

Descriptive statistics were used to summarise characteristics and clinical outcomes for the low, medium and high exposure groups in Morwell (tertiles of mean PM_2.5_ exposure in Morwell) as well as the non-exposed Sale group. Crude statistical significance was assessed using Pearson chi-squared tests for categorical and t-tests for continuous measures. Multivariate linear regression models were used to analyse the association between mean and peak PM_2.5_ exposure (controlling for key confounders) on continuous clinical outcomes. Logistic regression models were used for binary clinical outcomes. Oversampling of the asthmatic participants was corrected using the weighting method. Since smoking is the strongest risk factor for developing COPD, we tested whether the associations between PM_2.5_ exposure and outcomes differed between smoking groups by including an interaction between exposure and participants’ smoking status (non-smoker, ex-smoker and current smoker) for all the outcome variables. Whether there was an overall interaction effect was tested using a Wald test of all interaction terms equal to zero. Sensitivity analyses were performed with unweighted and complete case models. Multiple imputation by chained equation (MICE) was used to deal with missing data. Statistical analyses were performed using Stata version 15 (Stata Corporation, College Station, Texas 2015).

### Ethical considerations

The Monash University Human Research Ethics Committee (MUHREC) approved the Hazelwood Health Study: Cardiovascular and Respiratory Streams (approval number 1078). All participants provided written informed consent.

## RESULTS

As shown in Figure 1, 519 (39%) adults participated in the Respiratory Stream clinics; 346 from Morwell and 173 from Sale. Their characteristics were comparable across exposed groups (tertiles) and the unexposed group in terms of age, ethnicity, highest educational qualification, smoking status, years of smoking (for current and ex-smokers), occupational exposure and respiratory medication use in last 3 months, see Table 1. Distributions of exposure level for each group are provided in Supplementary Figure S1. However, there were higher proportions of male and obese participants and a lower proportion of employed participants in the higher exposure group compared with lower exposure group. Table 2 displays lung function and respiratory symptoms for participants in exposure groups. There were no substantial differences in post-bronchodilator spirometry or T_L_co between groups. However, respiratory symptoms in the last 12 months, including wheezing without an upper respiratory tract infection (URTI), chest tightness and chronic cough, were more prevalent in the higher exposure groups.

**Table 1.** Participant characteristics by exposure groups (tertiles of mean PM_2.5_ exposure in Morwell vs. Sale)

| Characteristics | Sale<br>(no exposure)<br>N=173 | Morwell low<br>exposure<br>N=109 | Morwell medium<br>exposure<br>N=113 | Morwell high<br>exposure<br>N=124 | p-value |
| --- | --- | --- | --- | --- | --- |
| **Age group, n (weighted %)** |  |  |  |  |  |
| 18-44 years | 44 (22%) | 36 (26%) | 36 (30%) | 35 (25%) | 0.74 |
| 45-64 | 74 (42%) | 43 (44%) | 43 (37%) | 50 (37%) |  |
| 65+ | 55 (36%) | 30 (29%) | 34 (34%) | 39 (38%) |  |
| **Age, weighted mean (SD)** | 57.3 (20.0) | 54.7 (14.3) | 54.5 (15.3) | 56.7 (14.7) | 0.50 |
| **Male, n (weighted %)** | 62 (36%) | 43 (46%) | 46 (43%) | 62 (56%) | 0.023 |
| **Caucasian / White, n (weighted %)** | 171 (99%) | 108 (99%) | 112 (100%) | 123 (99%) | 0.92 |
| **Employment status, n (weighted %)** |  |  |  |  |  |
| Employed | 89 (47%) | 44 (38%) | 46 (40%) | 50 (35%) | 0.021 |
| Retired | 57 (38%) | 32 (32%) | 33 (30%) | 37 (37%) |  |
| Unable to work | 10 (6%) | 15 (16%) | 8 (5%) | 15 (12%) |  |
| Other (unemployed, studying, home duties and other) | 17 (9%) | 18 (13%) | 26 (24%) | 22 (16%) |  |
| **Highest educational qualification, n (weighted %)** |  |  |  |  |  |
| Secondary up to year 10 | 34 (17%) | 28 (21%) | 29 (22%) | 28 (19%) | 0.79 |
| Secondary year 11-12 | 31 (19%) | 24 (23%) | 27 (24%) | 21 (17%) |  |
| Certificate (trade/ apprenticeship/ technical) | 73 (42%) | 34 (37%) | 38 (37%) | 53 (48%) |  |
| University or other Tertiary Institute degree | 34 (22%) | 22 (19%) | 16 (17%) | 21 (16%) |  |
| **BMI, n (weighted %)** |  |  |  |  |  |
| Underweight/Normal(BMI<25 kg/m<sup>2</sup>) | 40 (24%) | 23 (20%) | 21 (18%) | 15 (11%) | 0.06 |
| Overweight (25≤BMI<30 kg/m<sup>2</sup>) | 66 (38%) | 35 (33%) | 31 (29%) | 35 (30%) |  |
| Obese (BMI≥30 kg/m<sup>2</sup>) | 67 (38%) | 51 (47%) | 61 (53%) | 74 (59%) |  |
| **Smoking status, n (weighted %)** |  |  |  |  |  |
| Non-smoker | 82 (49%) | 58 (52%) | 60 (54%) | 49 (36%) | 0.10 |
| Ex-smoker | 66 (39%) | 35 (33%) | 34 (33%) | 51 (47%) |  |
| Current smoker | 25 (12%) | 16 (15%) | 19 (13%) | 24 (17%) |  |
| **Years of smoking among current and ex-smokers (N=270), weighted mean (SD)** | 23.6 (19.2) | 22.0 (11.0) | 20.5 (13.1) | 20.2 (12.3) | 0.54 |
| **Occupational exposure, n (weighted %)** | 64 (37%) | 43 (44%) | 44 (39%) | 54 (46%) | 0.47 |
| **Use any SAMA/SABA\* in the last 3 months, n (weighted %)** | 64 (31%) | 51 (33%) | 53 (36%) | 60 (36%) | 0.82 |
| **LAMA/LABA and/or ICS† use in the last 3 months, n (weighted %)** | 9 (4%) | 7 (3%) | 9 (6%) | 8 (6%) | 0.87 |

\* SAMA=short-acting muscarinic agonist, SABA=short-acting beta-agonist.
† LAMA=long-acting muscarinic agonist, LABA=long-acting beta-agonist, ICS=inhaled corticosteroids

**Table 2.** Lung function and respiratory symptoms by exposure groups (tertiles of mean PM_2.5_ exposure in Morwell vs. Sale)

| Outcome variables | Sale<br>(no exposure)<br>N=173 | Morwell low exposure<br>N=109 | Morwell medium<br>exposure<br>N=113 | Morwell high exposure<br>N=124 | p-value |
| --- | --- | --- | --- | --- | --- |
|  | n (weighted%) | n (weighted%) | n (weighted%) | n (weighted%) |  |
| **Spirometric COPD** | 24 (14%) | 9 (6%) | 14 (9%) | 12 (9%) | 0.27 |
| **Abnormal T<sub>L</sub>co** | 18 (11%) | 9 (7%) | 15 (10%) | 14 (12%) | 0.50 |
| **Spirometric COPD & abnormal T<sub>L</sub>co** | 7 (4%) | 2 (1%) | 6 (4%) | 6 (5%) | 0.27 |
|  | Weighted mean (SD) | Weighted mean (SD) | Weighted mean (SD) | Weighted mean (SD) |  |
| **Spirometry - post BD** |  |  |  |  |  |
| FEV<sub>1</sub>, Litres | 2.9 (1.1) | 3.0 (0.8) | 2.9 (0.8) | 2.9 (0.7) | 0.86 |
| FEV<sub>1</sub> z score | 0.0 (1.5) | 0.0 (1.0) | -0.3 (1.1) | -0.2 (1.0) | 0.22 |
| FEV<sub>1</sub> %Predicted | 99.3 (22.2) | 99.5 (13.9) | 95.9 (16.3) | 97.0 (14.9) | 0.29 |
| FVC, Litres | 3.7 (1.3) | 3.8 (1.1) | 3.7 (1.0) | 3.8 (0.8) | 0.94 |
| FVC z score | 0.2 (1.2) | 0.0 (0.9) | -0.1 (0.9) | -0.1 (0.8) | 0.10 |
| FVC %Predicted | 102.6 (18.4) | 100.7 (13.3) | 98.7 (13.5) | 99.2 (12.0) | 0.13 |
| FEV<sub>1</sub>/FVC | 76.5 (12.7) | 78.7 (7.0) | 77.2 (9.2) | 77.0 (8.5) | 0.20 |
| FEV<sub>1</sub>/FVC z score | -0.3 (1.4) | -0.1 (0.9) | -0.3 (1.0) | -0.3 (1.0) | 0.48 |
| **T<sub>L</sub>co** |  |  |  |  |  |
| T<sub>L</sub>co z score | 0.0 (1.7) | -0.2 (1.0) | -0.1 (1.1) | -0.1 (1.3) | 0.71 |
| T<sub>L</sub>co %Predicted | 101.0 (26.0) | 98.2 (15.0) | 99.1 (17.4) | 99.2 (19.9) | 0.68 |
| T<sub>L</sub>co Hb corrected z score | 0.0 (1.7) | -0.1 (0.9) | -0.1 (1.1) | -0.1 (1.2) | 0.73 |
| T<sub>L</sub>co Hb corrected %Predicted | 102.0 (24.9) | 98.8 (14.3) | 99.6 (17.3) | 99.8 (19.1) | 0.55 |
| **Respiratory symptoms in the last 12months** |  |  |  |  |  |
| Wheeze | 66 (34%) | 53 (41%) | 52 (38%) | 71 (50%) | 0.08 |
| Wheeze and breathlessness | 46 (23%) | 43 (30%) | 36 (25%) | 46 (31%) | 0.47 |
| Wheeze without URTI | 49 (23%) | 38 (27%) | 36 (23%) | 61 (43%) | <0.001 |
| Chest tightness | 28 (15%) | 22 (15%) | 30 (19%) | 36 (27%) | 0.036 |
| Dyspnoea at rest | 26 (13%) | 20 (16%) | 22 (14%) | 28 (19%) | 0.60 |
| Dyspnoea after exercise | 76 (42%) | 65 (54%) | 49 (38%) | 68 (52%) | 0.049 |
| Woken with dyspnoea | 12 (6%) | 17 (12%) | 20 (14%) | 16 (10%) | 0.20 |
| Woken with cough | 56 (29%) | 52 (45%) | 42 (34%) | 54 (39%) | 0.11 |
| Chronic cough | 55 (29%) | 47 (39%) | 52 (45%) | 67 (48%) | 0.040 |
| Chronic phlegm | 25 (13%) | 14 (13%) | 21 (16%) | 28 (22%) | 0.22 |

Adjusted associations between PM_2.5_ exposure, respiratory symptoms and lung function are displayed in Table 3. A 10 μg/m^3^ increase in mean PM_2.5_ exposure was associated with a 30% (95%CI: 3% to 64%) increase in the odds of chest tightness in the previous 12 months as well as a 24% (95%CI: 2% to 51%) increase in odds of chronic cough. Dose-response relationships between PM_2.5_ exposure and post BD spirometry and T_L_co z scores were not observed.

**Table 3.** Adjusted associations (Odds Ratios and 95% Confidence Intervals) between PM_2.5_ exposure, respiratory symptoms and lung function

|  | Mean exposure model<br>(10 µg/m<sup>3</sup>) |  | Peak exposure model<br>(100 µg/m<sup>3</sup>) |  |
| --- | --- | --- | --- | --- |
|  | Adj OR (95% CI) | p-value | Adj OR (95% CI) | p-value |
| **Respiratory symptoms in the last 12 months\*** |  |  |  |  |
| Wheeze | 1.13 (0.89,1.43) | 0.31 | 1.00 (0.89,1.13) | 0.99 |
| Wheeze and breathlessness | 1.00 (0.76,1.32) | 1.00 | 0.92 (0.80,1.06) | 0.26 |
| Wheeze without URTI | 1.19 (0.96,1.48) | 0.11 | 1.07 (0.96,1.20) | 0.22 |
| Chest tightness | 1.30 (1.03,1.64) | 0.026 | 1.08 (0.96,1.21) | 0.22 |
| Dyspnoea at rest | 1.02 (0.77,1.33) | 0.91 | 1.02 (0.87,1.19) | 0.84 |
| Dyspnoea after exercise | 1.08 (0.87,1.34) | 0.48 | 0.99 (0.89,1.11) | 0.88 |
| Woken with dyspnoea | 0.84 (0.63,1.12) | 0.23 | 0.92 (0.78,1.08) | 0.29 |
| Woken with cough | 0.97 (0.77,1.22) | 0.78 | 0.95 (0.84,1.07) | 0.39 |
| Chronic cough | 1.24 (1.02,1.51) | 0.035 | 1.06 (0.95,1.18) | 0.33 |
| Chronic phlegm | 1.23 (0.93,1.62) | 0.15 | 1.02 (0.88,1.17) | 0.83 |
| **Lung function†** |  |  |  |  |
| Spirometric COPD | 0.92 (0.67,1.26) | 0.60 | 1.02 (0.86,1.20) | 0.82 |
| Abnormal T<sub>lco</sub> | 1.18 (0.84,1.65) | 0.35 | 1.05 (0.89,1.24) | 0.55 |
|  | Adj β (95% CI) | p-value | Adj β (95% CI) | p-value |
| Post BD FEV<sub>1</sub> z score | 0.02 (-0.09,0.13) | 0.68 | 0.03 (-0.03,0.08) | 0.30 |
| Post BD FEV<sub>1</sub>/FVC z score | 0.04 (-0.06,0.13) | 0.44 | 0.00 (-0.05,0.05) | 0.99 |
| Corrected T<sub>lco</sub> z score | -0.05 (-0.18,0.08) | 0.48 | 0.02 (-0.05,0.09) | 0.60 |

\* Adjusted for smoking status, location of the participant (Morwell vs. Sale), BMI category, occupational exposure, nasal allergies/hayfever, employed or not and having a certificate, university or other tertiary institute degrees.
† Adjusted for smoking status, location of the participant (Morwell vs. Sale), BMI category, employed or not and having a certificate, university or other tertiary institute degrees.

Figure 2 presents the results from logistic regression models for respiratory symptoms when interactions between exposure and smoking status were included. For ease of interpretation, we report the estimated OR associated with exposure (a 10 μg/m^3^ increase in mean PM_2.5_ or a 100 μg/m^3^ increase in peak PM_2.5_ exposure) in each smoking group separately. A dose-response relationship existed between 10 μg/m^3^ increases in mean PM_2.5_ and chest tightness in the previous 12 months in non-smokers (OR 1.46, 95% CI 1.07, 2.00) and current smokers (OR 1.70, 95% CI 1.05, 2.75), but not in ex-smokers. For chronic cough, there was strong evidence for a dose-response relationship in current smokers (OR 2.13, 95%CI 1.24, 3.65 for each 10 μg/m^3^ increase in mean PM_2.5_. There was also some evidence suggesting a dose response relationship among non-smokers for wheeze without URTI in the previous 12 months and each 10 μg/m^3^ increase in mean PM_2.5_ (see Figure 2 and Table S1 in Supplementary Material).

**Figure 2.**
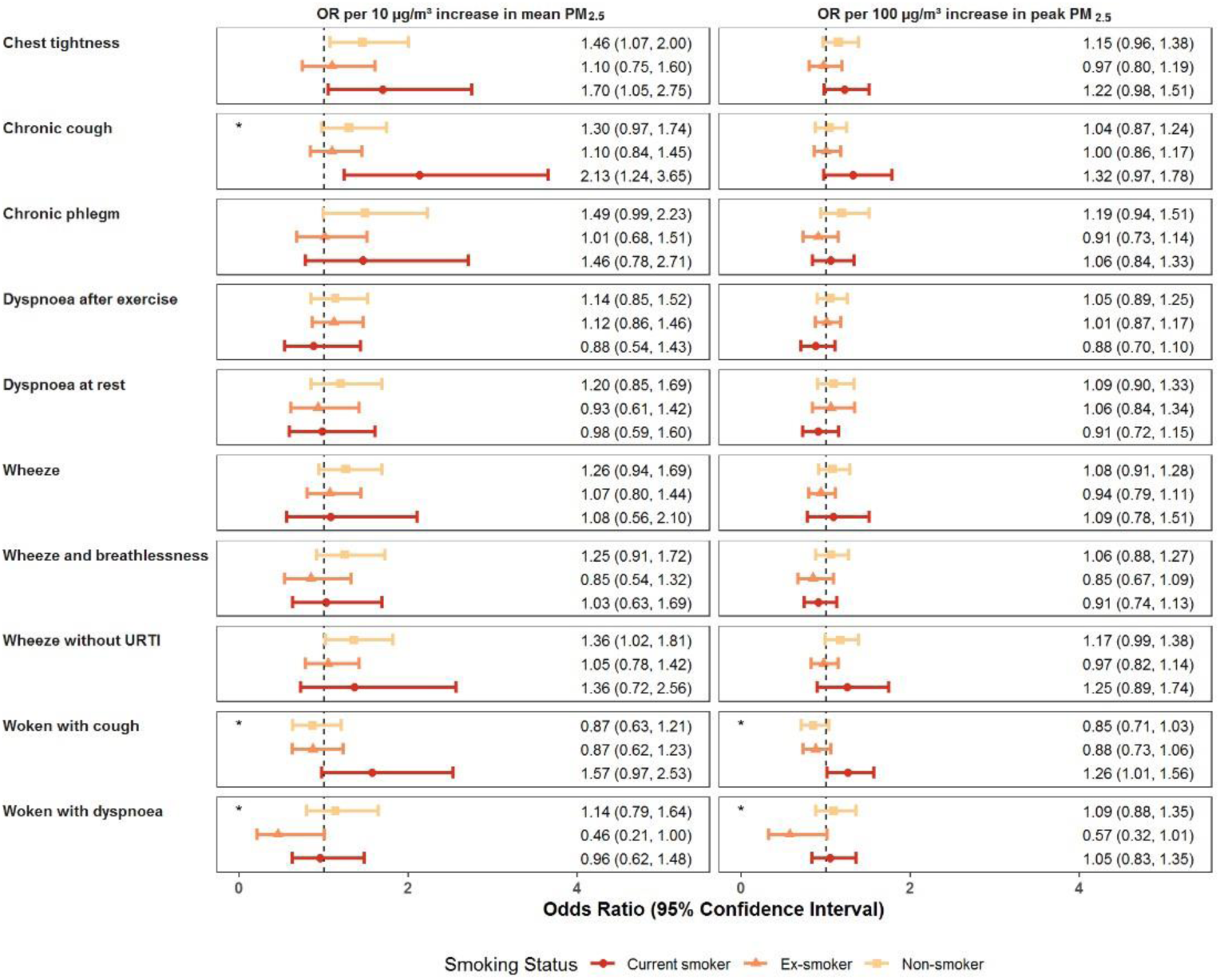
Forest plots of the estimated effects of PM_2.5_ exposure on respiratory symptoms in the past 12 months for each smoking group. Note: The OR were estimated using multivariate logistic regression with an interaction between exposure and smoking status. All models were also adjusted for age, gender, location of the participant (Morwell vs. Sale), BMI category, occupational exposure, nasal allergies/hayfever, employed or not and having a certificate university or other tertiary institute degrees. * indicates evidence of an overall interaction effect (p-value <0.1 from Wald test of all interaction terms equal to zero).

Figure 3 presents the results from regression models for lung function when interactions between exposure and smoking status were included. Among non-smokers, the interaction models estimated a 69% (95% CI: 11%, 158%) increase in the odds of spirometric COPD, a 52% increase in the odds of abnormal T_L_co (95% CI: 0%, 131%) and a 0.13 unit (95% CI: 0.00, 0.26) reduction in post BD FEV_1_/FVC z score per 10 µg/m^3^ increment in mean exposure to PM_2.5._ Among non-smokers, associations were also observed between 100 µg/m^3^ increments in peak exposure to PM_2.5._ and increased odds of Spirometric COPD, decreased BD FEV_1_/FVC z score and weaker evidence of increased odds of abnormal T_L_co. Evidence of interaction effects between exposure and smoking status were present for spirometric COPD and post BD FEV_1_/FVC z score but not abnormal T_L_co (see Figure 3 and Table S2 in the Supplementary Material).

**Figure 3.**
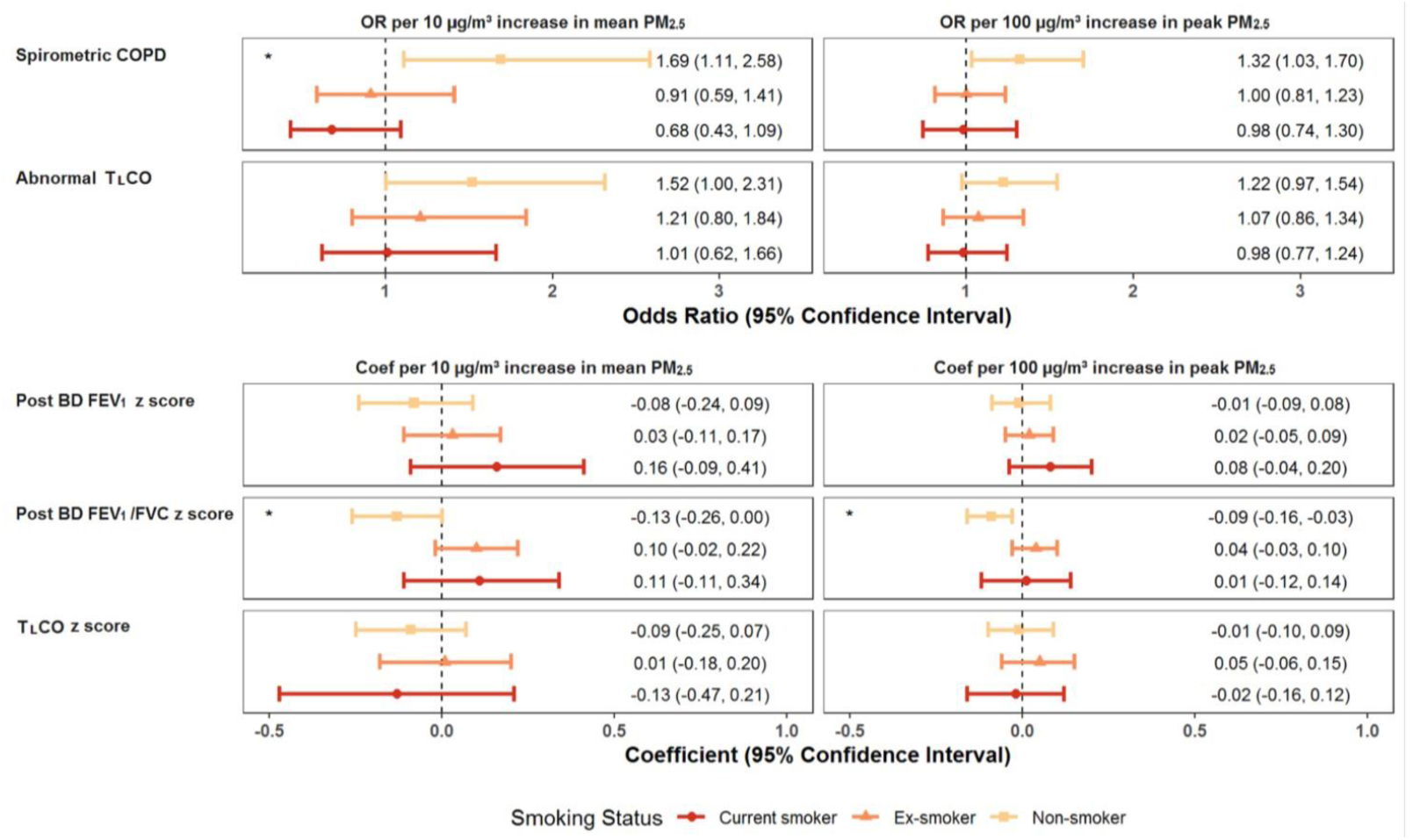
Forest plots of the estimated effect of PM_2.5_ exposure on lung function for each smoking group. Note: TLCO values are corrected for haemoglobin level. The ORs and coefficients were estimated using multivariate logistic and linear regression models with an interaction between exposure and smoking status. All models were adjusted for location of the participant (Morwell vs. Sale), BMI category occupational exposure, employment and higher education. * indicates evidence of an overall interaction effect (p-value <0.1 from Wald test of all interaction terms equal to zero).

## DISCUSSION

This analysis found that COPD, as defined by a post bronchodilator FEV_1_/FVC ratio less than the lower limit of normal, was associated with both mean and peak PM_2.5_ exposure from a coal mine fire in non-smoking individuals 3.5 to 4 years after exposure. In all participants, an association was also observed between mean mine-fire related PM_2.5_ exposure and respiratory symptoms (chest tightness and chronic cough) consistent with airflow obstruction. Effect modification by smoking was apparent for the associations between respiratory symptoms and PM_2.5_ exposure. This was clearest for chronic cough in the previous 12 months, with increments in mean PM_2.5_ exposure associated with a doubling of the odds of this symptom in current smokers.

The exposure-response relationship between PM2.5 and spirometric COPD in non-smokers in this study was consistent with findings in the study of Huang et al,[33] who investigated the association between COPD and PM_2.5_ in Taiwanese non-smokers. The relationship was also seen in a Taiwanese study by Guo et al[34] and a Chinese study from Zhao et al[35] who found PM_2.5_ exposure was associated with increased risk of COPD. Further, Zhao et al[35] suggested that PM_2.5_ and cigarette smoking exposure may have a “synergistic effect” in the development and progression of COPD. It is not clear why a similar association was not seen in current and ex-smokers in this study. It may be due to small numbers of participants in the current and ex-smoker in this study cohort or that the length of PM_2.5_ exposure in this study was weeks to months compared to years in the Zhao study.

The lack of an effect on spirometry for smokers and ex-smokers exposed to mine fire PM_2.5_ may equally relate to concurrent inhaled medication use, with these individuals potentially more likely to have used such medications compared to non-smokers. Lung function studies of asthmatics after a wildfire event have suggested a counter-intuitive preservation of lung function compared to non-asthmatics, with the possibility of a protective effect from the use of medications in this population.[8] Studies of COPD in non-smokers suggest a lower prevalence of respiratory symptoms, including chronic cough, reducing the likelihood of medication-seeking in this population.[36]

From an epidemiological perspective, a quarter to a third of COPD occurs in non-smokers, with the proportion greater in low-middle compared to high income countries.[36] This may relate to an important role for indoor air pollution in the pathogenesis of COPD in non-smokers, with 45% of non-smokers reported to be exposed to biomass and 73% to coal smoke.[36]

In general, there are limited data on the effects of particulate matter from coal mine fires on spirometry and our study provides a valuable addition to knowledge. Whilst comparison to other studies of coal mine fires is difficult for this reason, the similar emission profile between forest and coal mine fires allows comparison with the landscape fire literature.[1] Studies on the health impacts of wildfire smoke have found increasing evidence of a link with COPD using data on hospitalisations, physician visits and reliever medications.[8] However there were little data on effects on spirometry, with the exception of declines in peak expiratory flow among non-asthmatic children associated with wildfire exposure.[8] There was also little literature available on wild-fire smoke exposure lasting for up to six weeks, like the Hazelwood mine fire exposure. Wildfires often tend to be episodic and short in duration, and exposed populations from individual events are often small.[8] However, climate change may be contributing to increasing incidence of catastrophic wildfires globally, such as the recent megafires in Australia which burned over a six month period and exposed more than 10 million people.[37]

Declines in lung function (particularly FEV_1_) after smoke exposure are commonly reported from studies of cross shift and cross season variations in wildfire fire fighters.[37] One study reported an association between levoglucosan (a woodsmoke marker) and decline in FEV_1_ among wildland firefighters.[38] However, another study found no association between a decline in lung function and smoke components such as PM_3.5_, acrolein, formaldehyde or carbon monoxide exposure.[39]

The role of wildfire smoke causing sustained changes in lung function among adult fire fighters is unclear with contradictory results from existing studies.[37] In some children with asthma, peak expiratory flow rates have been measured to assess response to wildfires.[8] One small study measured FEV_1_ and peak flow prior to, and 1 month after, wildfires with high PM_2.5_ levels and found stable lung function, but also elevated sputum eosinophils in two subjects tested during the fires, suggesting an acute inflammatory effect.[40]

### Strengths & Limitations

A major strength of our study was the measurement of lung function through spirometry, to enable an objective definition of COPD. To our knowledge, there were no comparable data available from other populations exposed to coal mine fires. Wildfire studies have largely focused on hospital and primary care presentations or medication dispensing.[8] This stream achieved the required sample size. The findings build upon previously reported HHS respiratory health findings which have utilised administrative health datasets[16-18] and self-reported symptoms.[19]

Not relying on participants recalling their smoke exposure levels, an additional strength of our study was the estimation of individual exposures to PM_2.5_ from a combination of detailed time-location diaries and spatially and temporally resolved modelled PM_2.5_ concentrations using a chemical transport model. Whilst it is possible that some participants may have had difficulty recalling their precise locations and dates more than two years after the fire, considerable effort was made to manually review any detected inconsistencies in participant’s time-location diaries.

However, a limitation of the study includes that it was essentially a cross-sectional analysis and it was not possible to imply causation between PM_2.5_ exposure and COPD in non-smokers.

## Conclusions

Our study found an association in non-smokers exposed to PM_2.5_ from an open cut brown coal mine fire and increased risk of spirometry defined COPD almost four years after exposure Conversely, we also found a pronounced association between respiratory symptoms and PM_2.5_ exposure in smokers. These findings have important public health implications, as a better understanding of the exposure-response relationships across a range of PM_2.5_ exposure levels and durations would help inform policy decisions including evidence-based exposure reduction strategies, and long-term health service needs, for smoke-exposed communities. These may be particularly relevant to communities exposed to large-scale landscape fires such as those recently in Australia and the United States, and to future landscape fire events Our study also highlighted the importance of objective measurement of pulmonary impairment through spirometry, given that reliance on symptoms alone does not reliably identify disease, particularly in non-smokers. Long term followup of this exposed cohort may provide valuable insights into the incidence and prognosis of COPD in coal-mine fire PM_2.5_ exposed non-smokers.

## Data Availability

The Intellectual Property arising from the Hazelwood Health Study is owned by the Victorian Department of Health & Human Services. Any researchers wishing to collaborate with the Hazelwood Health Study researchers should contact the Principal Investigator

## ACKNOWLEDGMENTS

We gratefully acknowledge the contribution of all participating community members. We also thank Susan Denny, Kylie Sawyer, Shantelle Allgood and Kristina Thomas from the Monash University School of Rural Health who assisted with recruitment and data collection.

## CONTRIBUTORSHIP

SP led the drafting of the work and revising it critically, and contributed to interpretation of the data. BB contributed to the design of the work, the acquisition, analysis and interpretation of data, drafting the work and revising it critically. CG and JB contributed to the analysis and interpretation of data, drafting the work and revising it critically. DB contributed to the acquisition of data, drafting the work and revising it critically. JI contributed to the design of the work, the acquisition and interpretation of data, drafting the work and revising it critically. AM and TM contributed to the design of the work, the acquisition and interpretation of data, drafting the work and revising it critically. RH and BT contributed to the conception and design of the work, the interpretation of data, drafting the work and revising it critically. MA contributed to the conception and design of the work, the acquisition, analysis and interpretation of data, drafting the work and revising it critically. All authors approved the final version and agreed to be accountable for all aspects of the work in ensuring that questions related to the accuracy or integrity of any part of the work were appropriately investigated and resolved.

