## Supplementary Material for "Chronic Obstructive Pulmonary Disease in adults exposed to fine particles from a coal mine fire"

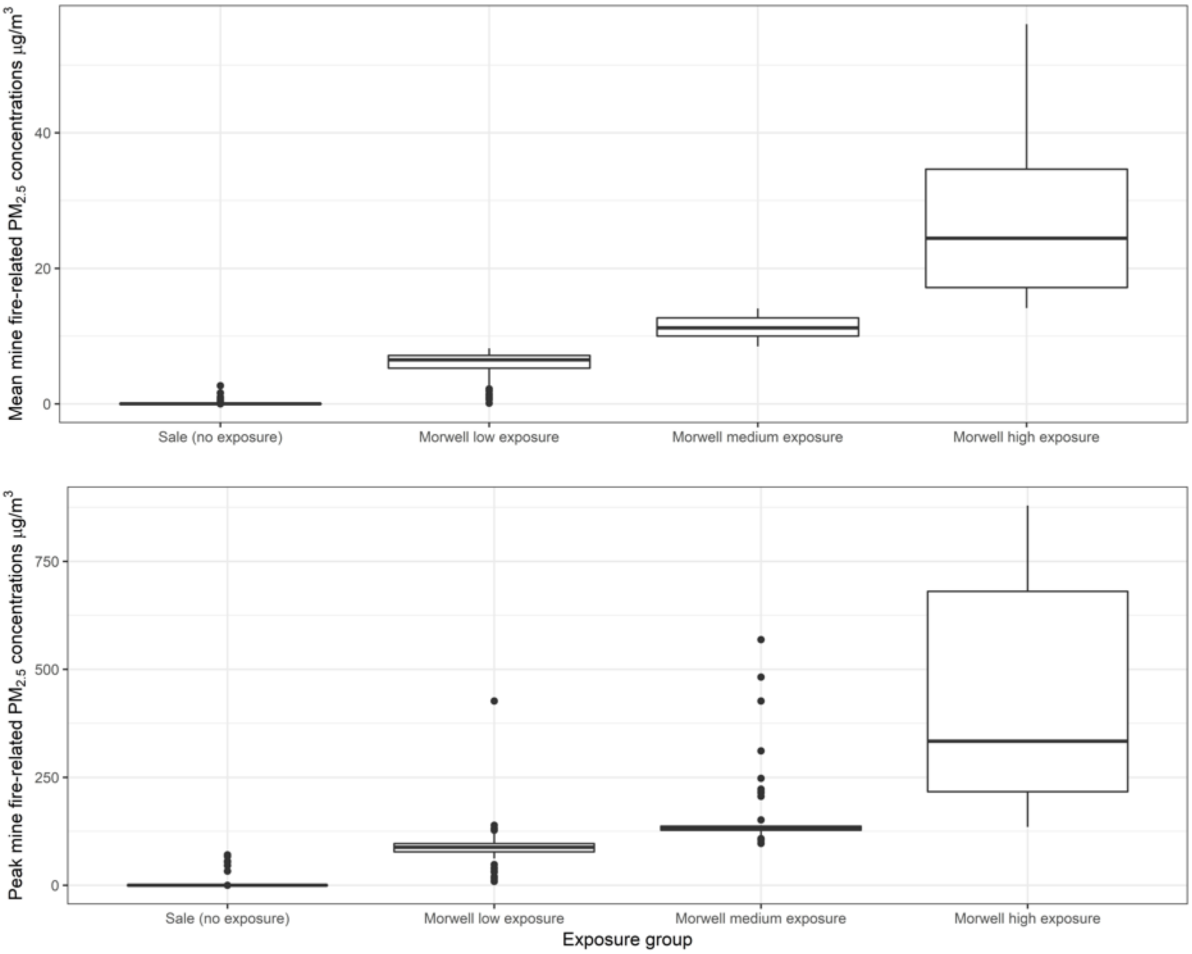


Figure S1. Box plot of mean and peak $PM_{2.5}$ exposure by exposure group (tertiles of mean $PM_{2.5}$ exposure in Morwell vs. Sale)

Table S1. Estimated effect sizes of $PM_{2.5}$ on respiratory symptoms by smoking status from the logistic models with interactions

| | Mean exposure (10 μg/m³) | | Peak exposure (100 μg/m³) | |
|---|---|---|---|---|
| | Adj OR* (95% CI) | p-value† | Adj OR* (95% CI) | p-value† |
| **Wheeze in the last 12 months** | | **0.68** | | **0.44** |
| Non-smoker | 1.26 (0.94, 1.69) | 0.12 | 1.08 (0.91, 1.28) | 0.38 |
| Ex-smoker | 1.07 (0.80, 1.44) | 0.65 | 0.94 (0.79, 1.11) | 0.46 |
| Current smoker | 1.08 (0.56, 2.10) | 0.82 | 1.09 (0.78, 1.51) | 0.62 |
| **Wheeze and breathlessness** | | 0.28 | | 0.30 |
| Non-smoker | 1.25 (0.91, 1.72) | 0.17 | 1.06 (0.88, 1.27) | 0.56 |
| Ex-smoker | 0.85 (0.54, 1.32) | 0.47 | 0.85 (0.67, 1.09) | 0.21 |
| Current smoker | 1.03 (0.63, 1.69) | 0.91 | 0.91 (0.74, 1.13) | 0.39 |
| **Wheeze without URTI** | | **0.40** | | **0.16** |
| Non-smoker | 1.36 (1.02, 1.81) | 0.034 | 1.17 (0.99, 1.38) | 0.06 |
| Ex-smoker | 1.05 (0.78, 1.42) | 0.73 | 0.97 (0.82, 1.14) | 0.69 |
| Current smoker | 1.36 (0.72, 2.57) | 0.34 | 1.25 (0.89, 1.75) | 0.20 |
| **Chest tightness in last 12 months** | | **0.28** | | **0.27** |
| Non-smoker | 1.46 (1.07, 1.99) | 0.016 | 1.15 (0.96, 1.38) | 0.14 |
| Ex-smoker | 1.10 (0.75, 1.60) | 0.62 | 0.97 (0.80, 1.19) | 0.80 |
| Current smoker | 1.70 (1.05, 2.75) | 0.032 | 1.22 (0.98, 1.51) | 0.07 |
| **Dyspnoea at rest in last 12 months** | | **0.58** | | **0.43** |
| Non-smoker | 1.20 (0.85, 1.68) | 0.31 | 1.09 (0.90, 1.33) | 0.38 |
| Ex-smoker | 0.93 (0.61, 1.43) | 0.75 | 1.06 (0.84, 1.34) | 0.64 |
| Current smoker | 0.97 (0.59, 1.60) | 0.92 | 0.91 (0.72, 1.14) | 0.40 |
| **Dyspnoea after exercise in last 12 months** | | **0.62** | | **0.40** |
| Non-smoker | 1.14 (0.85, 1.52) | 0.37 | 1.05 (0.89, 1.25) | 0.54 |
| Ex-smoker | 1.12 (0.86, 1.46) | 0.40 | 1.01 (0.87, 1.17) | 0.90 |
| Current smoker | 0.88 (0.54, 1.42) | 0.60 | 0.88 (0.70, 1.10) | 0.25 |
| **Woken with dyspnoea in last 12 months** | | **0.08** | | **0.09** |
| Non-smoker | 1.14 (0.79, 1.64) | 0.49 | 1.09 (0.88, 1.35) | 0.42 |
| Ex-smoker | 0.46 (0.21, 1.00) | 0.05 | 0.57 (0.32, 1.01) | 0.05 |
| Current smoker | 0.96 (0.62, 1.49) | 0.86 | 1.05 (0.83, 1.35) | 0.67 |
| **Woken with cough in last 12 months** | | **0.08** | | **0.014** |
| Non-smoker | 0.87 (0.63, 1.21) | 0.40 | 0.85 (0.71, 1.03) | 0.09 |
| Ex-smoker | 0.87 (0.62, 1.23) | 0.44 | 0.88 (0.73, 1.06) | 0.18 |
| Current smoker | 1.56 (0.97, 2.53) | 0.07 | 1.26 (1.01, 1.56) | 0.039 |
| **Chronic cough in the last 12 months** | | **0.07** | | **0.26** |
| Non-smoker | 1.30 (0.97, 1.75) | 0.08 | 1.04 (0.87, 1.24) | 0.69 |
| Ex-smoker | 1.10 (0.84, 1.45) | 0.48 | 1.00 (0.86, 1.17) | 0.97 |
| Current smoker | 2.18 (1.28, 3.71) | 0.004 | 1.32 (0.97, 1.79) | 0.07 |
| **Chronic phlegm in the last 12 months** | | **0.30** | | **0.24** |
| Non-smoker | 1.49 (0.99, 2.23) | 0.05 | 1.19 (0.94, 1.51) | 0.15 |
| Ex-smoker | 1.01 (0.68, 1.51) | 0.95 | 0.91 (0.73, 1.14) | 0.42 |
| Current smoker | 1.46 (0.78, 2.71) | 0.23 | 1.06 (0.84, 1.33) | 0.62 |

* OR estimated from logistic regression with an interaction between $PM_{2.5}$ exposure and smoking status. All models were also adjusted for location of the participant (Morwell vs. Sale), BMI category, occupational exposure, nasal allergies/hayfever, employment and having a higher education.

†P-value in line with individual OR represents the significant test for OR differed from 1 and **bold** p-value above represented p-value from Wald test of all interaction terms equal to zero, which indicated whether there was an overall significant interaction ($p<0.1$ suggests some evidence of an interaction effect).

Table S2. Estimated effect sizes of $PM_{2.5}$ on lung function by smoking status from the logistic or linear regression models with interactions.

| | Mean exposure (10 µg/m³) | | Peak exposure (100 µg/m³) | |
|---|---|---|---|---|
| | **Adj OR[*] (95% CI)** | **p-value‡** | **Adj OR[*] (95% CI)** | **p-value‡** |
| **Spirometric COPD** | | **0.008** | | **0.13** |
| Non-smoker | 1.69 (1.11, 2.58) | 0.015 | 1.32 (1.03, 1.70) | 0.029 |
| Ex-smoker | 0.91 (0.59, 1.41) | 0.68 | 1.00 (0.81, 1.23) | 0.99 |
| Current smoker | 0.68 (0.43, 1.09) | 0.11 | 0.98 (0.74, 1.30) | 0.89 |
| **Abnormal $T_Lco$** | | **0.39** | | **0.38** |
| Non-smoker | 1.52 ( 1.00, 2.31) | 0.048 | 1.22 ( 0.97, 1.54) | 0.09 |
| Ex-smoker | 1.21 ( 0.80, 1.84) | 0.36 | 1.07 ( 0.86, 1.34) | 0.52 |
| Current smoker | 1.01 ( 0.62, 1.66) | 0.96 | 0.98 ( 0.77, 1.24) | 0.86 |
| | **Adj β[†] (95%CI)** | **p-value‡** | **Adj β[†] (95%CI)** | **p-value‡** |
| **Post BD $FEV_1$ z score** | | **0.16** | | **0.43** |
| Non-smoker | -0.08 (-0.24, 0.09) | 0.35 | -0.01 (-0.09, 0.08) | 0.91 |
| Ex-smoker | 0.03 (-0.11, 0.17) | 0.67 | 0.02 (-0.05, 0.09) | 0.53 |
| Current smoker | 0.18 (-0.05, 0.42) | 0.12 | 0.09 (-0.03, 0.20) | 0.13 |
| **Post BD $FEV_1$/FVC z score** | | **0.005** | | **0.005** |
| Non-smoker | -0.13 (-0.26, -0.01) | 0.041 | -0.09 (-0.16, -0.03) | 0.004 |
| Ex-smoker | 0.11 (-0.01, 0.23) | 0.07 | 0.04 (-0.02, 0.11) | 0.19 |
| Current smoker | 0.12 (-0.11, 0.36) | 0.30 | 0.01 (-0.12, 0.15) | 0.87 |
| **$T_Lco$ z score** | | **0.60** | | **0.63** |
| Non-smoker | -0.09 (-0.25, 0.07) | 0.26 | -0.01 (-0.10, 0.09) | 0.86 |
| Ex-smoker | 0.01 (-0.18, 0.20) | 0.91 | 0.05 (-0.06, 0.15) | 0.36 |
| Current smoker | -0.13 (-0.47, 0.21) | 0.44 | -0.02 (-0.16, 0.12) | 0.76 |

[*] OR estimated from logistic regression including an interaction between $PM_{2.5}$ exposure and smoking status. All models were also adjusted for location of the participant (Morwell vs. Sale), BMI category, employed or not and having a certificate, university or other tertiary institute degrees.

[†] Coefficients estimated from linear regression including an interaction between $PM_{2.5}$ exposure and smoking status. All models were also adjusted for location of the participant (Morwell vs. Sale), BMI, employment and higher education

[‡] P-value in line with individual OR represents the significant test for OR differs from 1 and bold p-value above represents p-value from Wald test of all interaction terms equal to zero, which indicates whether there is an overall significant interaction (p<0.1 suggests some evidence of an interaction effect).
